# Fibrosis-4 Index and Long-Term Cardiovascular Outcomes in Primary Aldosteronism

**DOI:** 10.64898/2026.06.25.26356630

**Authors:** Cheng-Hsuan Tsai, Yu-Ching Chang, Chin-Chen Chang, Wan-Chen Wu, Yi-Yao Chang, Uei-Lin Chen, Bo-Ching Lee, Chi-Sheng Hung, Kuo-How Huang, Jeff Shih-Chieh Chueh, Vin-Cent Wu, Yen-Hung Lin

## Abstract

**Background:** The fibrosis-4 index (FIB-4) is a simple noninvasive marker originally developed to assess liver fibrosis risk. Accumulating evidence suggests that FIB-4 is associated with adverse cardiovascular outcomes, but its prognostic relevance in patients with primary aldosteronism (PA) remains unclear.

**Methods:** In this retrospective multicenter cohort study, patients with PA were stratified into low, intermediate, and high FIB-4 groups using established cutoffs: <1.3, 1.3–2.67, and >2.67. Outcomes included all-cause mortality, ischemic stroke, hemorrhagic stroke, acute myocardial infarction, and major adverse cardiovascular events (MACE). Multivariable Cox proportional hazards models were used to estimate hazard ratios (HRs) across FIB-4 categories.

**Results:** Among 2,467 patients with PA, 1,215 (49.3%) had FIB-4 <1.3, 863 (35.0%) had FIB-4 1.3–2.67, and 389 (15.8%) had FIB-4 >2.67. Patients with higher FIB-4 were older and had lower body mass index, worse renal function, lower potassium, and a higher comorbidity burden. During follow-up, all-cause mortality increased across FIB-4 categories, from 13.9% in the low FIB-4 group to 58.1% in the high FIB-4 group. After multivariable adjustment, FIB-4 >2.67 was associated with higher risks of all-cause mortality (adjusted HR, 1.98; 95% CI, 1.54–2.54) and MACE (adjusted HR, 1.78; 95% CI, 1.44–2.20), compared with FIB-4 <1.3. Adjusted linear regression analyses showed that higher FIB-4 was significantly associated with lower renin and higher aldosterone-to-renin ratio.

**Conclusions:** In patients with PA, elevated FIB-4 identified a high-risk subgroup with increased all-cause mortality and MACE. FIB-4 may serve as a simple, noninvasive tool for prognostic stratification in patients with PA.

**Novelty and Relevance:** *What Is New?:*

- In this retrospective multicenter cohort of patients with primary aldosteronism, elevated FIB-4 identified a high-risk subgroup with increased all-cause mortality and MACE.
- FIB-4 was associated with PA-related biochemical markers, including plasma renin activity and the aldosterone-to-renin ratio, suggesting a link between FIB-4 and renin suppression in PA.
- This is the first study to evaluate the prognostic significance of FIB-4 in patients with PA and to examine its relationship with PA-related biochemical features.

*What Is Relevant?:*

- PA is clinically heterogeneous, and conventional biochemical markers may not fully capture the cumulative multisystem risk burden in high-risk patients.
- FIB-4 is a simple, noninvasive, and routinely available index that may complement current risk stratification in PA by integrating biochemical, hepatic-metabolic, renal, and comorbidity-related information.

*Clinical Implications?:*

- Incorporating FIB-4 into PA assessment may help identify patients who warrant more comprehensive evaluation of cardiovascular, renal, hepatic, and metabolic risk.
- Elevated FIB-4 may support closer clinical follow-up and more intensive risk-factor optimization in high-risk patients with PA.

## Introduction

Primary aldosteronism (PA) is an important cause of hypertension and is increasingly recognized as a major contributor to cardiovascular morbidity and mortality^1,2^. Beyond its effects on blood pressure, excess aldosterone promotes vascular inflammation, myocardial fibrosis, renal injury, and metabolic dysregulation, leading to a disproportionate burden of cardiovascular and renal complications compared with essential hypertension^3-5^. Despite advances in the diagnosis and treatment of PA, risk stratification remains challenging because patients with PA are clinically heterogeneous, ranging from relatively mild biochemical phenotypes to advanced multisystem disease with established target-organ damage^6-9^.

Liver-related metabolic dysfunction and fibrosis have been increasingly linked to adverse cardiovascular outcomes, including among patients with hypertension^10^. The fibrosis-4 index (FIB-4), calculated from age, aminotransferase levels, and platelet count, was originally developed as a noninvasive marker of liver fibrosis^11^. However, accumulating evidence suggests that FIB-4 may also serve as a marker of broader systemic vulnerability, with higher values associated with cardiovascular disease, cerebrovascular disease, chronic kidney disease, heart failure, and mortality in diverse populations^10,12-16^. By integrating age, hepatic injury, and platelet count, FIB-4 may reflect cumulative cardiometabolic, inflammatory, and fibrotic burden beyond that captured by conventional cardiovascular risk factors.

Whether FIB-4 provides prognostic information in patients with PA remains unknown. This question is clinically relevant because PA is characterized by aldosterone-mediated cardiovascular and renal injury, and a simple, routinely available, noninvasive index may help identify patients at particularly high long-term risk. Therefore, in this multicenter cohort study, we examined the association between FIB-4 and long-term clinical outcomes among patients with PA. We further explored whether FIB-4 was associated with biochemical markers of PA, including plasma renin activity, plasma aldosterone concentration, and the aldosterone-to-renin ratio (ARR).

## Methods

### Data Source

This retrospective multicenter cohort study used data from the National Taiwan University Hospital–integrated Medical Database, which includes electronic medical records from eight hospitals within the National Taiwan University Hospital health care system across Taiwan. The database contains structured information on demographics, diagnoses, laboratory measurements, prescriptions, procedures, and clinical outcomes, and is maintained using standardized data management procedures to ensure data accuracy and completeness. Disease diagnoses were identified using International Classification of Diseases, Ninth Revision, Clinical Modification codes before 2015 and International Classification of Diseases, Tenth Revision, Clinical Modification codes thereafter. This study was approved by the Research Ethics Committee of National Taiwan University Hospital.

### Study Population

We identified adult patients with hypertension who underwent testing for PA, defined as available measurements of both plasma renin activity and plasma aldosterone concentration, between January 1, 2006, and December 31, 2022. Hypertension was defined by diagnostic codes for hypertension and/or documented use of antihypertensive medications. For patients with multiple renin measurements during the study period, the first available plasma renin activity measurement was used as the index test.

PA was defined biochemically according to the 2025 Endocrine Society clinical practice guideline^17^. Patients were classified as having PA if they had suppressed renin, defined as plasma renin activity ≤1.0 ng/mL/h, together with plasma aldosterone concentration ≥10 ng/dL and an ARR >20 ng/dL per ng/mL/h. Plasma renin activity and plasma aldosterone concentration were measured using immunoassays as part of routine clinical care across participating hospitals. Patients younger than 18 years of age and those without available laboratory data required to calculate the fibrosis-4 index were excluded.

### Fibrosis-4 Index

The fibrosis-4 index was calculated at baseline using age, aspartate aminotransferase, alanine aminotransferase, and platelet count according to the following formula:

FIB-4 = age × AST / (platelet count × √ALT)

where age is expressed in years, AST and ALT are expressed in U/L, and platelet count is expressed as 10⁹/L. Patients were categorized into 3 groups using established clinical cutoffs: low FIB-4 (<1.3), intermediate FIB-4 (1.3–2.67), and high FIB-4 (>2.67). The low FIB-4 group was used as the reference group in outcome analyses. FIB-4 was also analyzed as a continuous variable in restricted cubic spline models to evaluate potential nonlinear associations with clinical outcomes.

### Covariates

Baseline characteristics were collected at the time of the index test, including age, sex, body mass index, body weight, body height, laboratory measurements, comorbidities, cardiovascular history, and antihypertensive medication use. Laboratory measurements included plasma renin activity, plasma aldosterone concentration, ARR, serum creatinine, estimated glomerular filtration rate, serum potassium, fasting blood glucose, low-density lipoprotein cholesterol, high-density lipoprotein cholesterol, triglycerides, uric acid, aspartate aminotransferase, alanine aminotransferase, and platelet count.

Comorbidities and cardiovascular history were identified using diagnosis codes and included diabetes mellitus, hyperlipidemia, coronary artery disease, chronic kidney disease, atrial fibrillation, heart failure, prior myocardial infarction, prior intracranial hemorrhage, and prior ischemic stroke. Baseline antihypertensive medication use included renin-angiotensin system inhibitors, beta blockers, alpha blockers, calcium channel blockers, diuretics, and mineralocorticoid receptor antagonists. The number of antihypertensive medication classes at baseline was also recorded.

### Outcomes

The main outcomes of interest were all-cause mortality and major adverse cardiovascular events (MACE). MACE was defined as a composite of ischemic stroke, intracranial hemorrhage, acute myocardial infarction, and all-cause mortality. Individual nonfatal cardiovascular outcomes, including ischemic stroke, intracranial hemorrhage, and acute myocardial infarction, were analyzed separately. All-cause mortality was ascertained through linkage with the National Death Registry. Cardiovascular events were identified using inpatient diagnosis codes.

### Statistical Analysis

Baseline characteristics were summarized according to FIB-4 categories: low FIB-4 (<1.3), intermediate FIB-4 (1.3–2.67), and high FIB-4 (>2.67). Continuous variables were presented as mean ± standard deviation or median with interquartile range, as appropriate, and categorical variables were presented as number and percentage. Trends across FIB-4 categories were evaluated using linear regression for continuous variables and the Cochran-Armitage trend test for binary categorical variables.

Incidence rates were calculated as the number of events per 100 person-years. Cox proportional hazards models were used to estimate hazard ratios (HR) and 95% confidence intervals (CIs) for the associations between FIB-4 categories and clinical outcomes, using FIB-4 <1.3 as the reference group. Patients were followed from the date of the index test until the occurrence of the outcome of interest, loss to follow-up, or the end of data availability, whichever occurred first. Multivariable Cox models were adjusted for age, sex, body mass index, estimated glomerular filtration rate, serum potassium, diabetes mellitus, chronic kidney disease, number of antihypertensive medications, renin-angiotensin system inhibitor use, beta blocker use, alpha blocker use, thiazide diuretic use, and loop diuretic use. FIB-4 was also analyzed as a continuous variable using restricted cubic spline models to evaluate potential nonlinear associations with all-cause mortality. FIB-4 was log-transformed before spline modeling, and restricted cubic splines were constructed with knots at the 5th, 35th, 65th, and 95th percentiles. HRs were plotted using FIB-4 = 1.3 as the reference value, with the x-axis presented in the original FIB-4 scale after back-transformation.

Multivariable linear regression models were used to examine the associations between FIB-4 and PA-related biochemical markers, including plasma renin activity, plasma aldosterone concentration, and the ARR. Log-transformed FIB-4 was modeled as the dependent variable, and each log-transformed biochemical marker was modeled separately as the independent variable. Models were adjusted for age, sex, body mass index, estimated glomerular filtration rate, serum potassium, diabetes mellitus, chronic kidney disease, number of antihypertensive medications, renin-angiotensin system inhibitor use, beta blocker use, alpha blocker use, thiazide diuretic use, and loop diuretic use. Adjusted association plots were generated for visualization by residualizing both log-transformed FIB-4 and each log-transformed biochemical marker against the same covariates. β coefficients and P values shown in the plots were derived from the corresponding multivariable linear regression models. Restricted cubic spline analyses and adjusted association plots were performed using R software, version 4.5.3, with the survival, rms, and ggplot2 packages. Other statistical analyses were performed using IBM SPSS Statistics, version 25.0. A two-sided P value <0.05 was considered statistically significant.

## Results

### Study Population

Among 7,378 hypertensive patients with available FIB-4, renin and aldosterone testing, 2,467 met the biochemical criteria for PA and were included in the final analytic cohort. Among these patients, 1,215 patients (49.3%) had low FIB-4 (<1.3), 863 (35.0%) had intermediate FIB-4 (1.3–2.67), and 389 (15.8%) had high FIB-4 (>2.67) (**Figure 1**).

**Figure 1.**
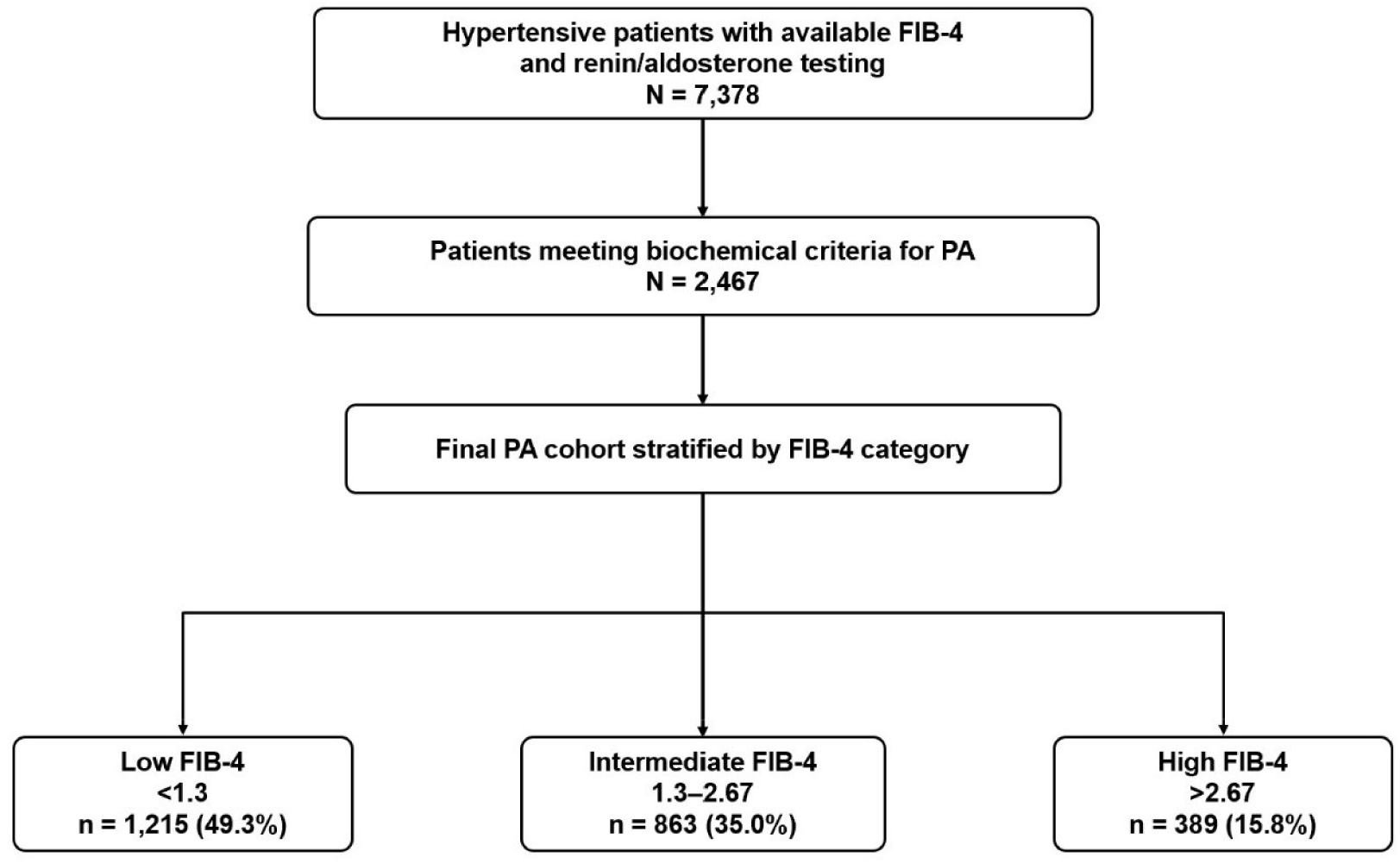
Study flowchart. Among 7,378 hypertensive patients with available FIB-4 and renin/aldosterone testing, 2,467 met the biochemical criteria for PA and were included in the final analytic cohort. Patients were stratified according to established FIB-4 categories. FIB-4 indicates fibrosis-4 index; PA, primary aldosteronism.

### Baseline Characteristics According to FIB-4 Categories

Baseline characteristics according to FIB-4 categories are shown in **Table 1**. Patients with higher FIB-4 were older, had lower body mass index, lower body weight, lower body height, worse renal function, and lower serum potassium levels. The mean age increased from 52.4 ± 13.4 years in the low FIB-4 group to 73.1 ± 12.6 years in the high FIB-4 group, and the proportion of patients aged 65 years or older increased from 16.8% to 75.8% across FIB-4 categories. Body mass index decreased progressively from 26.2 ± 4.7 kg/m² in the low FIB-4 group to 23.8 ± 4.1 kg/m² in the high FIB-4 group.

**Table 1.**
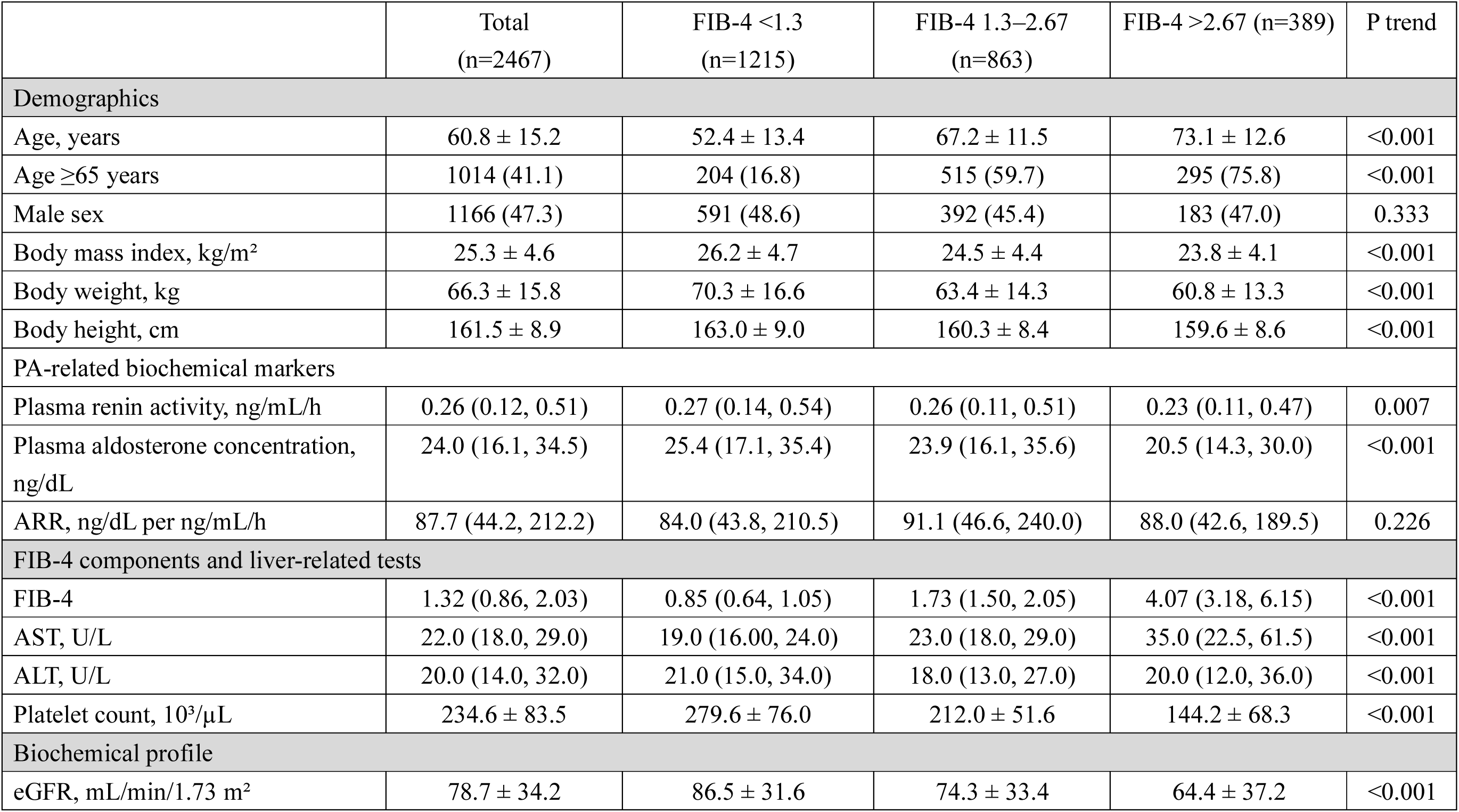
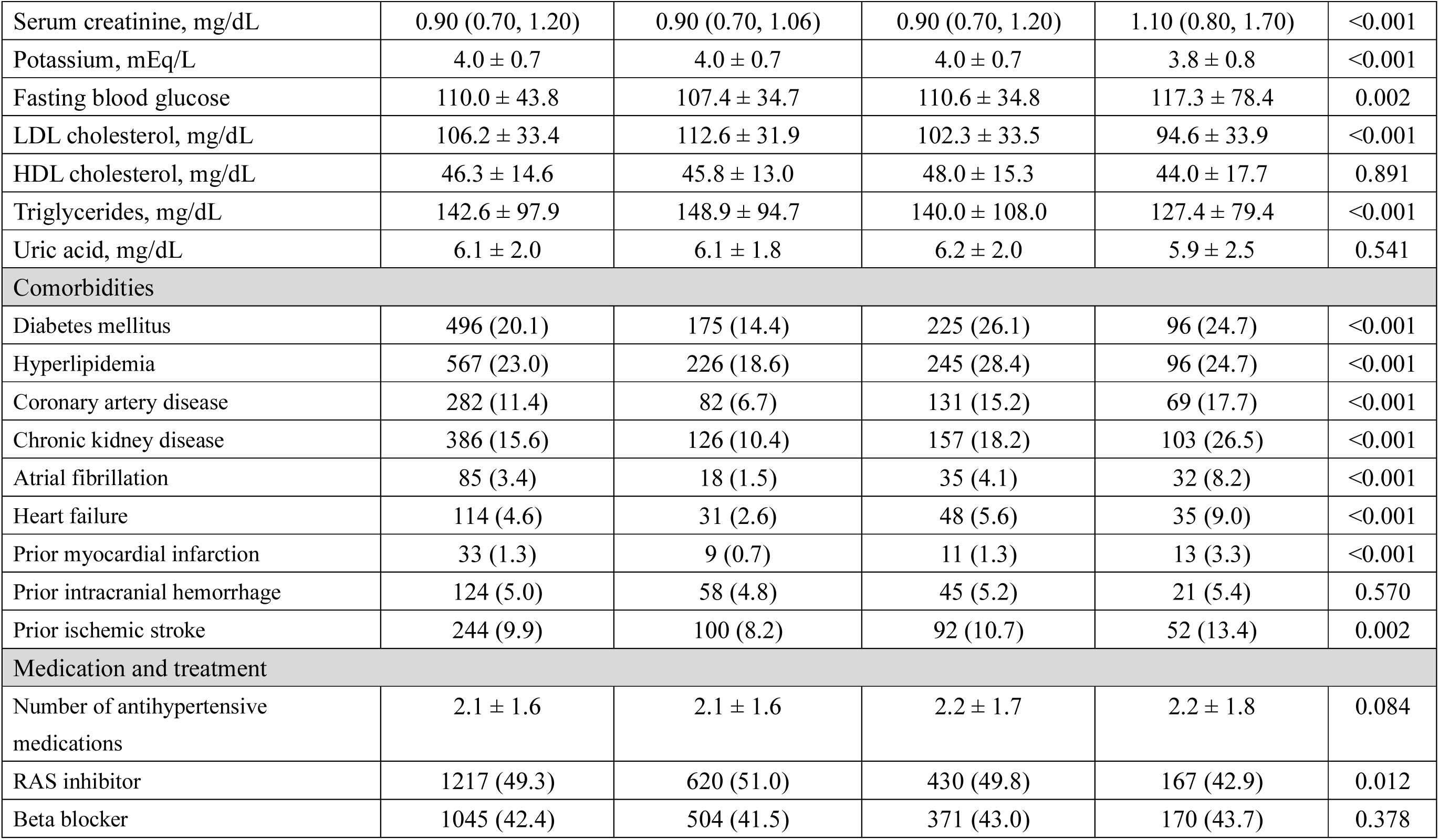
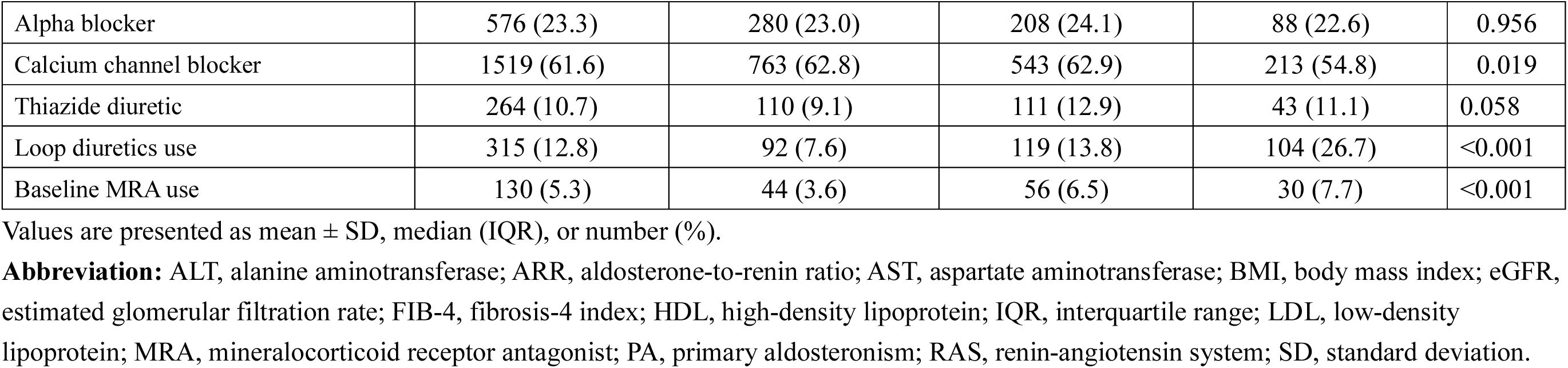
Baseline Characteristics of Patients with Primary Aldosteronism According to FIB-4 Categories.

PA-related biochemical markers differed modestly across FIB-4 categories. Plasma renin activity decreased slightly across increasing FIB-4 categories, from a median of 0.27 ng/mL/h in the low FIB-4 group to 0.23 ng/mL/h in the high FIB-4 group. Plasma aldosterone concentration also decreased across FIB-4 categories, from 25.4 ng/dL to 20.5 ng/dL. In contrast, the ARR did not show a significant trend across FIB-4 categories.

As expected, the components of FIB-4 differed substantially across categories. Patients with higher FIB-4 had higher AST levels and markedly lower platelet counts. The median FIB-4 increased from 0.85 in the low FIB-4 group to 4.07 in the high FIB-4 group. Higher FIB-4 was also associated with a higher burden of comorbidities, including diabetes mellitus, hyperlipidemia, coronary artery disease, chronic kidney disease, atrial fibrillation, heart failure, prior myocardial infarction, and prior ischemic stroke. Baseline loop diuretic and mineralocorticoid receptor antagonist use were also more common among patients with higher FIB-4.

### Clinical Outcomes According to FIB-4 Categories

Clinical outcomes according to FIB-4 categories are shown in **Table 2**. During a median follow-up of 3.88 years (IQR, 1.69–8.01), all-cause mortality increased substantially across FIB-4 categories. All-cause mortality occurred in 169 patients (13.9%) in the low FIB-4 group, 262 patients (30.4%) in the intermediate FIB-4 group, and 226 patients (58.1%) in the high FIB-4 group. The corresponding incidence rates were 2.51, 5.56, and 16.45 events per 100 person-years, respectively.

**Table 2.**
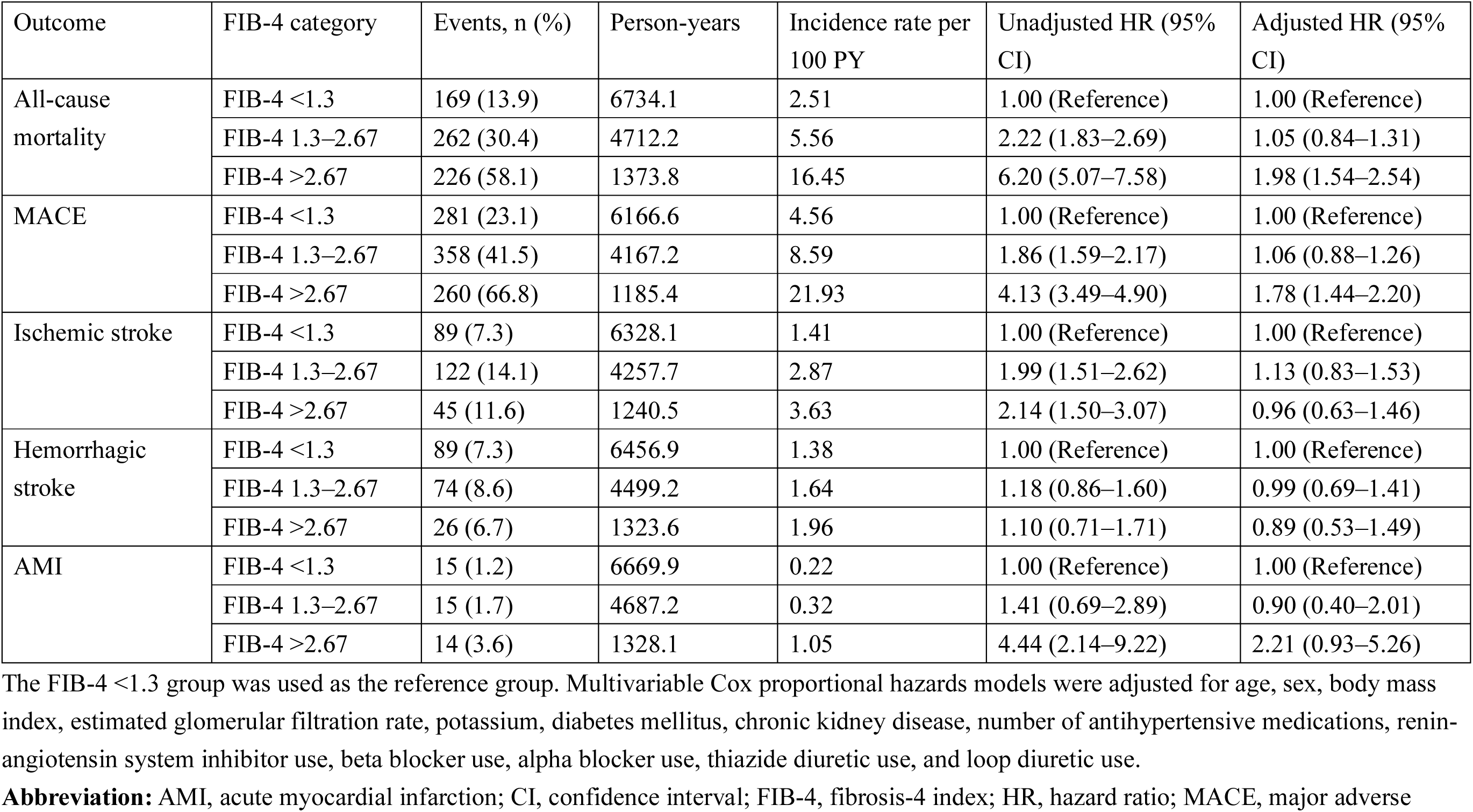
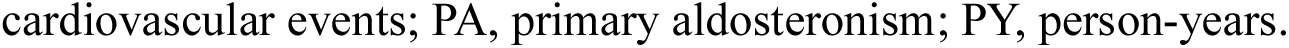
Clinical Outcomes According to FIB-4 Categories in Patients with Primary Aldosteronism.

| Outcome | FIB-4 category | Events, n (%) | Person-years | Incidence rate per 100 PY | Unadjusted HR (95% CI) | Adjusted HR (95% CI) |
| --- | --- | --- | --- | --- | --- | --- |
| All-cause mortality | FIB-4 <1.3 | 169 (13.9) | 6734.1 | 2.51 | 1.00 (Reference) | 1.00 (Reference) |
|  | FIB-4 1.3–2.67 | 262 (30.4) | 4712.2 | 5.56 | 2.22 (1.83–2.69) | 1.05 (0.84–1.31) |
|  | FIB-4 >2.67 | 226 (58.1) | 1373.8 | 16.45 | 6.20 (5.07–7.58) | 1.98 (1.54–2.54) |
| MACE | FIB-4 <1.3 | 281 (23.1) | 6166.6 | 4.56 | 1.00 (Reference) | 1.00 (Reference) |
|  | FIB-4 1.3–2.67 | 358 (41.5) | 4167.2 | 8.59 | 1.86 (1.59–2.17) | 1.06 (0.88–1.26) |
|  | FIB-4 >2.67 | 260 (66.8) | 1185.4 | 21.93 | 4.13 (3.49–4.90) | 1.78 (1.44–2.20) |
| Ischemic stroke | FIB-4 <1.3 | 89 (7.3) | 6328.1 | 1.41 | 1.00 (Reference) | 1.00 (Reference) |
|  | FIB-4 1.3–2.67 | 122 (14.1) | 4257.7 | 2.87 | 1.99 (1.51–2.62) | 1.13 (0.83–1.53) |
|  | FIB-4 >2.67 | 45 (11.6) | 1240.5 | 3.63 | 2.14 (1.50–3.07) | 0.96 (0.63–1.46) |
| Hemorrhagic stroke | FIB-4 <1.3 | 89 (7.3) | 6456.9 | 1.38 | 1.00 (Reference) | 1.00 (Reference) |
|  | FIB-4 1.3–2.67 | 74 (8.6) | 4499.2 | 1.64 | 1.18 (0.86–1.60) | 0.99 (0.69–1.41) |
|  | FIB-4 >2.67 | 26 (6.7) | 1323.6 | 1.96 | 1.10 (0.71–1.71) | 0.89 (0.53–1.49) |
| AMI | FIB-4 <1.3 | 15 (1.2) | 6669.9 | 0.22 | 1.00 (Reference) | 1.00 (Reference) |
|  | FIB-4 1.3–2.67 | 15 (1.7) | 4687.2 | 0.32 | 1.41 (0.69–2.89) | 0.90 (0.40–2.01) |
|  | FIB-4 >2.67 | 14 (3.6) | 1328.1 | 1.05 | 4.44 (2.14–9.22) | 2.21 (0.93–5.26) |

The FIB-4 <1.3 group was used as the reference group. Multivariable Cox proportional hazards models were adjusted for age, sex, body mass index, estimated glomerular filtration rate, potassium, diabetes mellitus, chronic kidney disease, number of antihypertensive medications, renin-angiotensin system inhibitor use, beta blocker use, alpha blocker use, thiazide diuretic use, and loop diuretic use.
**Abbreviation:** AMI, acute myocardial infarction; CI, confidence interval; FIB-4, fibrosis-4 index; HR, hazard ratio; MACE, major adverse
cardiovascular events; PA, primary aldosteronism; PY, person-years.

In unadjusted Cox models, both intermediate and high FIB-4 were associated with higher risks of all-cause mortality compared with low FIB-4. After multivariable adjustment, high FIB-4 remained independently associated with all-cause mortality, whereas intermediate FIB-4 was not. Compared with FIB-4 <1.3, FIB-4 >2.67 was associated with an adjusted HR of 1.98 (95% CI, 1.54–2.54) for all-cause mortality.

A similar pattern was observed for MACE. MACE occurred in 281 patients (23.1%) in the low FIB-4 group, 358 patients (41.5%) in the intermediate FIB-4 group, and 260 patients (66.8%) in the high FIB-4 group. The incidence rate of MACE increased from 4.56 per 100 person-years in the low FIB-4 group to 21.93 per 100 person-years in the high FIB-4 group. After multivariable adjustment, FIB-4 >2.67 remained significantly associated with a higher risk of MACE (adjusted HR, 1.78; 95% CI, 1.44–2.20), whereas FIB-4 1.3–2.67 was not significantly associated with MACE.

For individual cardiovascular outcomes, higher FIB-4 categories were associated with higher crude event rates for ischemic stroke and acute myocardial infarction. After multivariable adjustment, FIB-4 categories were not significantly associated with ischemic stroke or hemorrhagic stroke. For acute myocardial infarction, the adjusted association did not reach statistical significance, although the point estimate remained elevated in the high FIB-4 group (adjusted hazard ratio, 2.21; 95% CI, 0.93–5.26).

### Continuous Association Between FIB-4 and Outcomes

When FIB-4 was modeled as a continuous variable using restricted cubic splines, higher FIB-4 was associated with a progressively higher risk of all-cause mortality (**Figure 2A**). The association between FIB-4 and all-cause mortality was significant overall (P for overall association <0.001), with evidence of both a linear component (P for linearity <0.001) and nonlinearity (P for nonlinearity = 0.004). The risk increased above the clinical cutoff of 1.3 and rose more steeply at higher FIB-4 values. In categorical analyses, the high FIB-4 group consistently showed the strongest association with adverse outcomes (**Figure 2B**).

**Figure 2.**
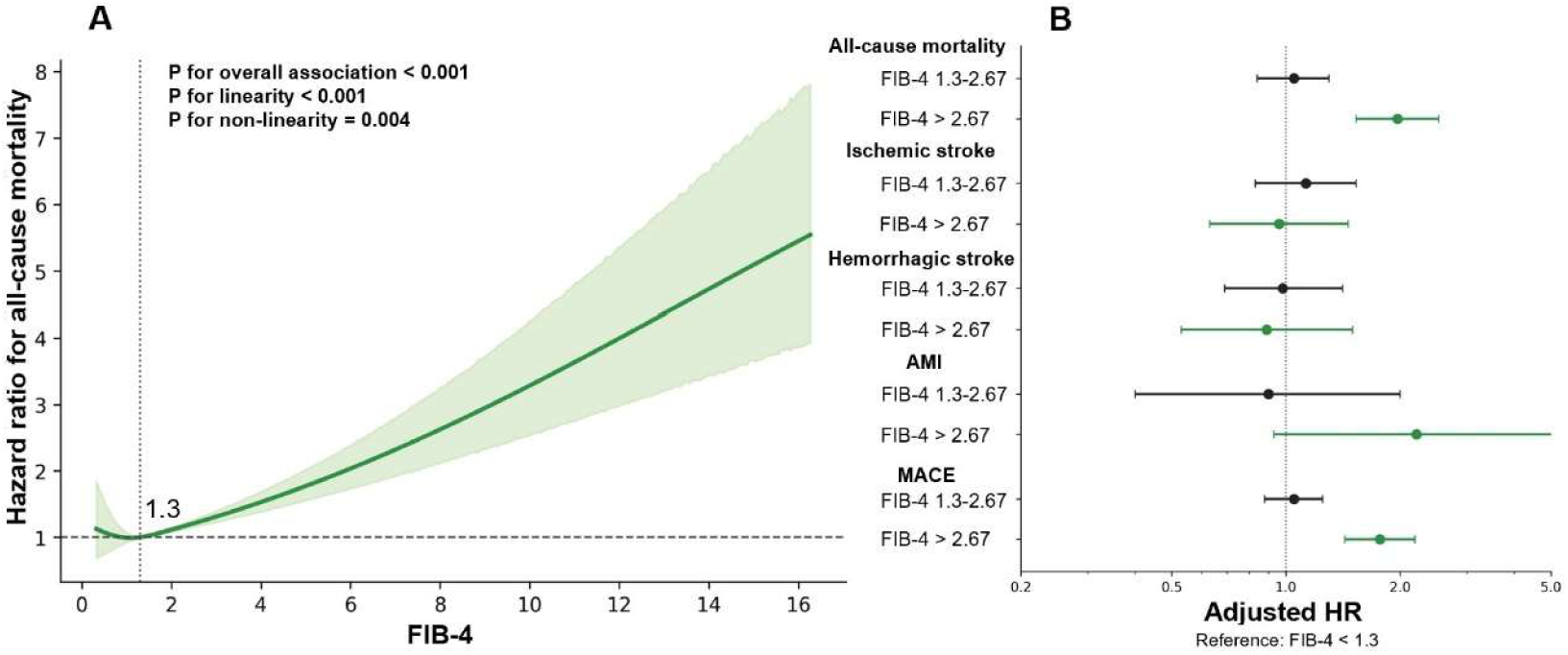
Association between FIB-4 and long-term clinical outcomes in patients with primary aldosteronism. (A) Restricted cubic spline analysis showing the adjusted association between FIB-4 and all-cause mortality, using FIB-4 = 1.3 as the reference value. The solid line indicates the adjusted HR, and the shaded area indicates the 95% confidence interval. (B) Adjusted HRs for clinical outcomes according to FIB-4 categories, using FIB-4 <1.3 as the reference group. AMI indicates acute myocardial infarction; CI, confidence interval; FIB-4, fibrosis-4 index; HR, hazard ratio; MACE, major adverse cardiovascular events; PA, primary aldosteronism.

### Associations Between FIB-4 and PA-Related Biochemical Markers

The associations between FIB-4 and PA-related biochemical markers are shown in **Figure 3**. In adjusted residualized association plots, higher log-transformed FIB-4 was modestly associated with lower log plasma renin activity (β = -0.0395, P = 0.0016). Plasma aldosterone concentration was not significantly associated with FIB-4 after multivariable adjustment (β = -0.0393, P = 0.1226). A modestly positive association was observed between log-transformed ARR and log-transformed FIB-4 (β = 0.0242, P = 0.0395).

**Figure 3.**
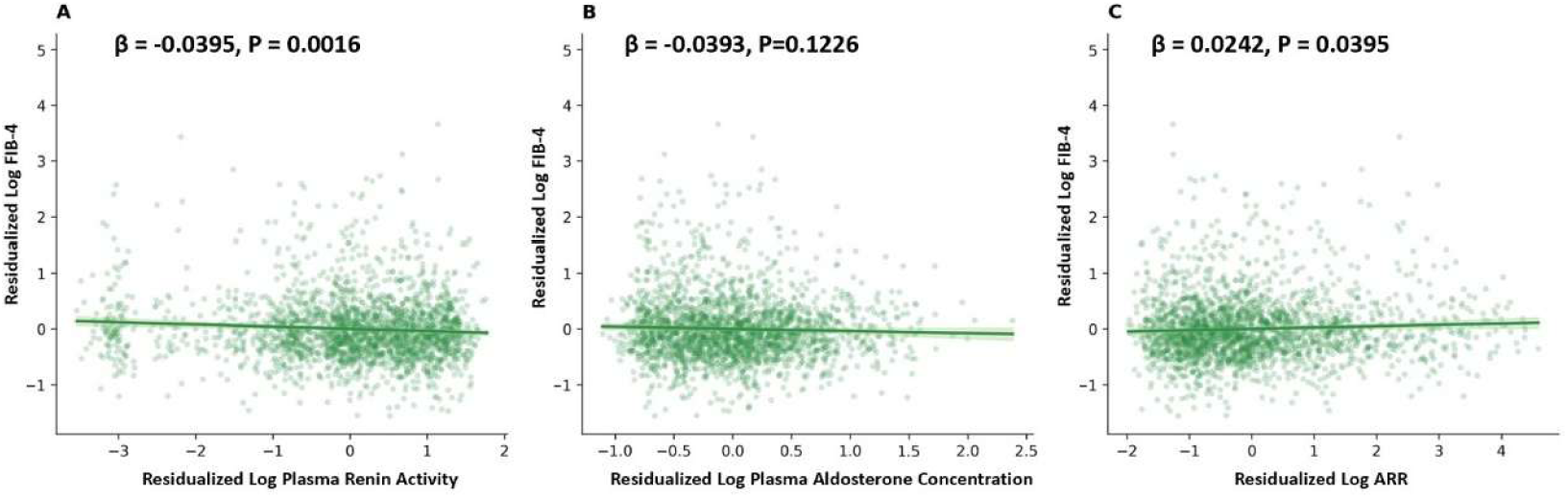
Associations Between FIB-4 and Primary Aldosteronism–Related Biochemical Markers. Adjusted residualized association plots showing the relationships between log-transformed FIB-4 and log-transformed plasma renin activity (A), plasma aldosterone concentration (B), and ARR (C). β coefficients and P values were derived from multivariable linear regression models. ARR indicates aldosterone-to-renin ratio; FIB-4, fibrosis-4 index; PA, primary aldosteronism.

## Discussion

In this multicenter cohort of patients with PA, elevated FIB-4 identified a clinically vulnerable subgroup with substantially higher risks of all-cause mortality and MACE. Patients with higher FIB-4 were older and had lower body mass index, impaired renal function, lower potassium levels, and a greater burden of cardiometabolic and cardiovascular comorbidities. Importantly, FIB-4 was also associated with plasma renin activity and ARR, suggesting a link between FIB-4 and renin suppression in patients with PA. Together, these findings support FIB-4 as an integrated prognostic marker that may aid risk stratification by capturing both PA-related biochemical features and broader multisystem risk burden.

Although FIB-4 was originally developed as a noninvasive marker to estimate liver fibrosis risk, its prognostic relevance may extend beyond liver disease alone11,18-22. By incorporating age, aminotransferase levels, and platelet count, FIB-4 may capture cumulative biological stress related to aging, hepatic-metabolic dysfunction, inflammation, platelet-related factors, and systemic illness. In the present study, higher FIB-4 was accompanied by worse renal function, lower potassium levels, and a higher prevalence of diabetes mellitus, coronary artery disease, chronic kidney disease, atrial fibrillation, heart failure, and prior ischemic stroke. This pattern is consistent with growing evidence that liver-related metabolic dysfunction is closely linked to cardiovascular risk. For example, in a large cohort of patients with steatotic liver disease, metabolic dysfunction-associated steatotic liver disease was associated with higher risks of new-onset heart failure, heart failure hospitalization, and heart failure with preserved ejection fraction14. Previous investigations have also reported that higher FIB-4 is associated with all-cause mortality, cardiovascular mortality, heart failure, cerebrovascular disease, and major adverse cardiovascular events across diverse clinical settings11,18,19,21-26. However, whether FIB-4 carries prognostic information in PA has not been previously established. To our knowledge, this is the first study to evaluate the prognostic significance of FIB-4 specifically among patients with PA and to further examine its relationship with PA-related biochemical markers.

Importantly, the observed associations between FIB-4 and PA-related biochemical markers provide additional insight into its potential relevance in PA. Higher FIB-4 was associated with lower plasma renin activity and higher ARR after multivariable adjustment, suggesting that FIB-4 may be linked to the renin-suppressed biochemical phenotype of PA. Higher FIB-4 was also accompanied by lower serum potassium levels, a clinical marker of more pronounced mineralocorticoid receptor activation and PA severity. These findings suggest that FIB-4 may integrate PA-related biochemical features with downstream organ damage and multisystem risk burden.

The mechanisms underlying the association between FIB-4 and renin suppression remain uncertain. One possible explanation is age-related dysregulation of aldosterone physiology27-29. Aging has been associated with decreased normal zona glomerulosa CYP11B2 expression and increased aldosterone-producing cell clusters, a histopathologic pattern that may contribute to age-related autonomous aldosteronism and renin suppression30. Because age is also a component of FIB-4, this age-related adrenal remodeling may partly explain the observed relationship between higher FIB-4 and a more renin-suppressed biochemical phenotype. However, FIB-4 likely reflects more than age alone, as it also incorporates aminotransferase levels and platelet count and was associated with comorbidity burden and adverse outcomes after multivariable adjustment, including adjustment for age.

These findings have potential implications for risk stratification in PA. Current clinical assessment of PA emphasizes blood pressure, biochemical severity, hypokalemia, renal function, cardiovascular comorbidities, and evidence of target-organ damage17,31-33. However, patients with PA are heterogeneous, and conventional biochemical markers may not fully capture the cumulative burden of systemic injury6,7,9,34-38. FIB-4 may complement existing clinical assessment by providing a readily available marker that integrates hepatic-metabolic stress, platelet-related vulnerability, renal dysfunction, and comorbidity burden. In this context, elevated FIB-4 may help identify a high-risk PA phenotype characterized by greater target-organ vulnerability and higher long-term mortality risk. Patients with PA and elevated FIB-4 may warrant more comprehensive assessment for cardiovascular, renal, and metabolic complications, closer longitudinal follow-up, and aggressive optimization of global cardiovascular risk factors. Importantly, FIB-4 should be interpreted as an adjunctive risk stratification marker rather than as a replacement for established PA evaluation or a standalone determinant of PA-specific treatment selection. Further studies are needed to determine whether FIB-4 can inform risk-based management strategies in PA, including among patients treated with aldosterone-targeted therapy.

Several limitations should be acknowledged. First, this was a retrospective observational study, and residual confounding cannot be excluded. Second, PA was defined biochemically using available renin and aldosterone measurements based on current guideline suggestions17, and aldosterone suppression testing, adrenal venous sampling, and subtype classification were not available. In addition, plasma renin activity and plasma aldosterone concentration were measured in routine clinical settings across multiple hospitals over a long study period; therefore, variability in assay platforms, calibration, reagents, and sampling conditions may have introduced measurement heterogeneity. Third, FIB-4 was used as a risk marker rather than as a definitive diagnosis of liver fibrosis; data on liver imaging, elastography, alcohol use, viral hepatitis, and other liver-specific etiologies were not systematically available.

Fourth, age is a component of FIB-4 and is also a strong determinant of mortality. Although age was included in multivariable models, residual age-related confounding may remain. Fifth, treatment decisions, including mineralocorticoid receptor antagonist therapy and adrenalectomy, were not randomized and may have influenced long-term outcomes. Finally, the study was conducted within a single health care system in Taiwan, and the generalizability of these findings to other populations and health care settings requires further studies.

### Perspectives

In conclusion, elevated FIB-4 identified a high-risk subgroup of patients with PA characterized by greater comorbidity burden and worse long-term cardiovascular outcomes. These findings support FIB-4 as a simple, noninvasive integrative marker for risk stratification in PA, potentially linking the renin-suppressed biochemical phenotype of PA with broader multisystem risk burden. Incorporating FIB-4 into clinical assessment may help identify patients who warrant more comprehensive evaluation of cardiovascular, renal, hepatic, and metabolic risks and subsequent more intensive risk-factor optimization to improve clinical outcomes.

## Nonstandard Abbreviation and Acronyms

ARR: aldosterone-to-renin ratio
CI: confidence interval
HR: hazard ratio
FIB-4: fibrosis-4 index
MACE: major adverse cardiovascular events
PA: primary aldosteronism

## Data availability

The data underlying this article will be shared on reasonable request to the corresponding author.

## Acknowledgements

We thank the staff of the Eighth and Second Core Lab in the Department of Medical Research at National Taiwan University Hospital for technical support. We also thank all members of the TAIPAI Study Group for help during the study. We would like to express our thanks to the staff of Department of Medical Research for providing clinical data from National Taiwan University Hospital-integrative Medical Data Center. The authors also thank Alfred Hsing-Fen Lin, Zoe Ya-Zhu Syu, and Winnie Su-Hui Wu, who served in Raising Statistics Consultant Inc., for their assistance with statistical analysis.

